# Predicting Conversion from Mild Cognitive Impairment to Alzheimer’s Disease Using a Vision Transformer and Hippocampal MRI Slices

**DOI:** 10.1101/2025.05.21.25328062

**Authors:** René Seiger, Peter Fierlinger, Alzheimer’s Disease Neuroimaging Initiative

**Affiliations:** Technical University of Munich, TUM School of Natural Sciences, Physics Department, Garching, Germany

## Abstract

Convolutional neural networks (CNNs) have been the standard for computer vision tasks including applications in Alzheimer’s disease (AD). Recently, Vision Transformers (ViTs) have been introduced, which have emerged as a strong alternative to CNNs. A common precursor stage of AD is a syndrome called mild cognitive impairment (MCI). However, not all individuals diagnosed with MCI progress to AD. In this investigation, we aimed to assess whether a ViT can reliably predict converters versus non-converters. A transfer learning approach was used for model training, by applying a pretrained ViT model, fine-tuned on the ADNI dataset. The cohort comprised 575 individuals (299 stable MCI; 276 progressive MCI who converted within 36 months), from whom axial T1-weighted MRI slices covering the hippocampal region were used as model input. Results showed an average area under the receiver operating characteristic curve (AUC-ROC) on the test set of 0.74 ± 0.02 (mean ± SD), an accuracy of 0.69 ± 0.03, a sensitivity of 0.65 ± 0.07, a specificity of 0.72 ± 0.06, and an F1-score for the progressive MCI class of 0.67 ± 0.04. These findings demonstrate that a ViT approach achieves reasonable classification accuracy for predicting the conversion from MCI to AD by specifically focusing on the hippocampal region.

## 1. Introduction

Convolutional neural networks (CNNs) [1,2] have long served as the main architecture for visual processing tasks and are widely used for classification and prediction based on magnetic resonance imaging (MRI) data. However, transformer models, introduced by Vaswani et al. [3] and utilizing the attention mechanism [4], have recently revolutionized the field of natural language processing (NLP) and have also proven valuable for vision tasks involving image data [5]. The Vision Transformer (ViT) architecture offers several advantages over classical CNN approaches, making it a formidable architecture for the medical domain, where MRI and similar imaging techniques are used to assess disease status and prognosis. While CNNs focus primarily on local or adjacent regions of an image, ViTs are capable of modeling contextualized long-range dependencies on a global scale via the self-attention mechanism. Images are divided into fixed-size patches, which are treated as input tokens, similar to how words are processed in classical Transformers used for natural language processing tasks. When trained on sufficiently large datasets, ViTs are able to learn more generalized and flexible representations, while, unlike CNNs, not relying on inductive biases such as locality or translation equivariance [5,6].

Hence, reliable, accurate, and well-performing deep learning models are needed, as they could serve as valuable tools with the potential to support physicians in the classification and prediction of various diseases, particularly Alzheimer’s disease (AD), where early diagnosis is key. AD is one of the most prevalent conditions among elderly individuals, with estimates indicating that over 30 million people worldwide are affected and numbers are steadily increasing [7]. To date, no cure exists, and its underlying causes remain poorly understood, although research indicates that brain atrophy patterns, which can be detected with MRI, are closely linked to, and likely result from, the pathological accumulation and spread of amyloid-beta and tau proteins [8]. The precursor stage of AD is referred to as mild cognitive impairment (MCI). During this phase, signs of cognitive decline particularly related to memory are already present, although activities of daily life are basically not impaired [9]. Estimates vary, but approximately 15% of individuals with MCI progress to Alzheimer’s dementia within two years, and about 33% do so within five years. Conversely, around 26% of individuals with MCI revert to normal cognition [10]. It has been shown in people with a rare genetic form of AD that neuropathological changes underlying dementia are already present in the brain several years before any clinical symptoms are evident [11]. This relatively long preclinical phase makes early detection using modern deep learning algorithms particularly valuable and promising. Therefore, developing accurate classification models to project the cognitive trajectory of individuals with MCI presents a critical opportunity to reduce patient suffering and long-term health care costs.

While ViTs have already demonstrated excellent performance in classifying AD patients from non-demented subjects based on MRI data [12,13] and can be considered a viable alternative to CNNs for this task, studies evaluating their predictive power regarding AD remain scarce. A study by Hoang et al. [14] has explored this question using structural MRI. However, following the methodological procedure reported in their investigation, data leakage likely occurred as previously noted by Valizadeh et al. [15]. Hence, results presented in their work probably represent overly optimistic accuracy metrics rather than an unbiased assessment of a ViT in predicting the conversion from MCI to AD. Therefore, further research is needed to determine whether this ViT approach provides state of the art results, as seen in CNNs for example, which was the dedicated objective of our present study.

To this end, three consecutive axial brain slices covering the hippocampal region were selected as input to our model. We focused on this area located in the temporal lobe, as it is one of the earliest brain structures affected by the disease [16] and is strongly associated with the processing of memory-related content and memory function [17]. Thus, by focusing on slices encompassing this region, we aim to capture early, subtle morphological changes that may be indicative of MCI-to-AD conversion. One drawback of the ViT architecture, however, is its need for massive training data [5], which poses a challenge in medical imaging, where available datasets typically include MRI scans from thousands of subjects at best. If not properly addressed, these models are prone to overfitting, memorizing the training data without generalizing to unseen cases. This issue can be mitigated by using pretrained models, which are already trained on millions of images and can be fine-tuned through a transfer learning approach using a specific dataset relevant to the target task. To account for this, we applied a ViT architecture which was pretrained on millions of images and fine-tuned it on a dataset of MRI slices for our specific task of MCI-to-AD prediction.

## 2. Methods

### 2.1. Magnetic Resonance Imaging Data

Data from the Alzheimer’s Disease Neuroimaging Initiative (ADNI) were used for this investigation. As stated by ADNI, the initiative was launched in 2003 as a public-private partnership, led by Principal Investigator Michael W. Weiner, MD. The original goal of ADNI was to test whether serial MRI, positron emission tomography (PET), other biological markers, and clinical and neuropsychological assessment can be combined to measure the progression of MCI and early AD. The current goals include validating biomarkers for clinical trials, improving the generalizability of ADNI data by increasing diversity in the participant cohort, and to provide data concerning the diagnosis and progression of AD to the scientific community. For up-to-date information, see adni.loni.usc.edu.

### 2.2. Demographics and MCI Status

The baseline MRI scan from participants with a diagnosis of MCI in ADNI1-3 and ADNIGO was included in the analysis. Subjects were divided according to their assessments into a stable (sMCI) and a progressive (pMCI) group based on the following approach: Individuals in the sMCI group were categorized as stable if they retained this status for a minimum of three years and maintained it throughout all available follow-up assessments provided by ADNI. The pMCI group included participants who progressed from MCI at baseline to Alzheimer’s disease (AD) within 36 months. Individuals who transitioned from MCI to AD and subsequently reverted to MCI were excluded. Similarly, participants who converted from MCI to a cognitively normal (CN) status, or who then reverted from CN back to MCI, were also excluded. In total, data from 575 participants were included, with 299 in the sMCI group and 276 in the pMCI group. Details regarding demographics and clinical assessment scores are provided in Table 1.

**Table 1.**
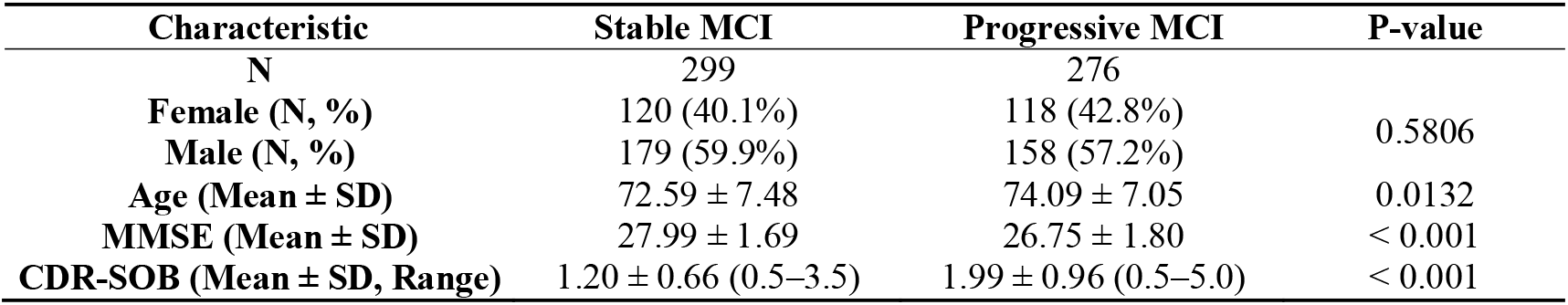
Demographic information and clinical assessment scale scores. MMSE: Mini-Mental State Examination; CDR-SOB: Clinical Dementia Rating – Sum of Boxes.

| Characteristic | Stable MCI | Progressive MCI | P-value |
| --- | --- | --- | --- |
| N | 299 | 276 |  |
| Female (N, %) | 120 (40.1%) | 118 (42.8%) | 0.5806 |
| Male (N, %) | 179 (59.9%) | 158 (57.2%) |  |
| Age (Mean ± SD) | 72.59 ± 7.48 | 74.09 ± 7.05 | 0.0132 |
| MMSE (Mean ± SD) | 27.99 ± 1.69 | 26.75 ± 1.80 | < 0.001 |
| CDR-SOB (Mean ± SD, Range) | 1.20 ± 0.66 (0.5–3.5) | 1.99 ± 0.96 (0.5–5.0) | < 0.001 |

### 2.3. MRI Data Preprocessing and Preparation

Structural 3D T1-weighted MRI data provided by ADNI were available either processed (for details see: https://adni.loni.usc.edu/data-samples/adni-data/neuroimaging/mri/mri-pre-processing/) or in raw DICOM format. Available DICOM scans were transformed into NIfTI format, while all data subsequently underwent the same preprocessing pipeline. The T1-weighted scans were processed using MATLAB (The MathWorks Inc., Natick, Massachusetts, USA) and the SPM12 toolbox (https://www.fil.ion.ucl.ac.uk/spm/software/spm12/, version 7771). Structural images underwent bias field correction to account for intensity inhomogeneities. Gray and white matter probability maps, thresholded at 0.5, were then used as masks to extract brain tissue from the bias-corrected volumes. Normalization to MNI standard space was performed via the forward deformation fields estimated during segmentation, resulting in one unified volume with dimensions 157 × 189 × 156 and a voxel size of 1 × 1 × 1 mm^3^. Three consecutive axial slices of the 3D volume of each subject centered around the hippocampal area were extracted (MNI space from z = −17 to z = −19) and assigned to the respective 2D RGB-image channels of the model for further analysis.

### 2.4. Model Architecture – Vision Transformer (ViT)

A ViT architecture ‘vit_base_patch16_224’ from the TIMM (PyTorch Image Models) library was used as the backbone for our experiments. The model was pretrained on ImageNet and natively processes 224 × 224 RGB images by dividing them into 16 × 16 non-overlapping patches. The original model included a classification head consisting of a single linear layer that maps the 768-dimensional transformer output to 1000 output classes, corresponding to the ImageNet dataset. To adapt this model for our task, we modified the classification head by inserting a dropout layer (p = 0.3) before the final linear layer to reduce overfitting, and adjusted the final output to produce a single value for binary classification. The original model architecture is described in [5], while the used ViT model with our custom adjustments are depicted in Figure 1.

**Figure 1.**
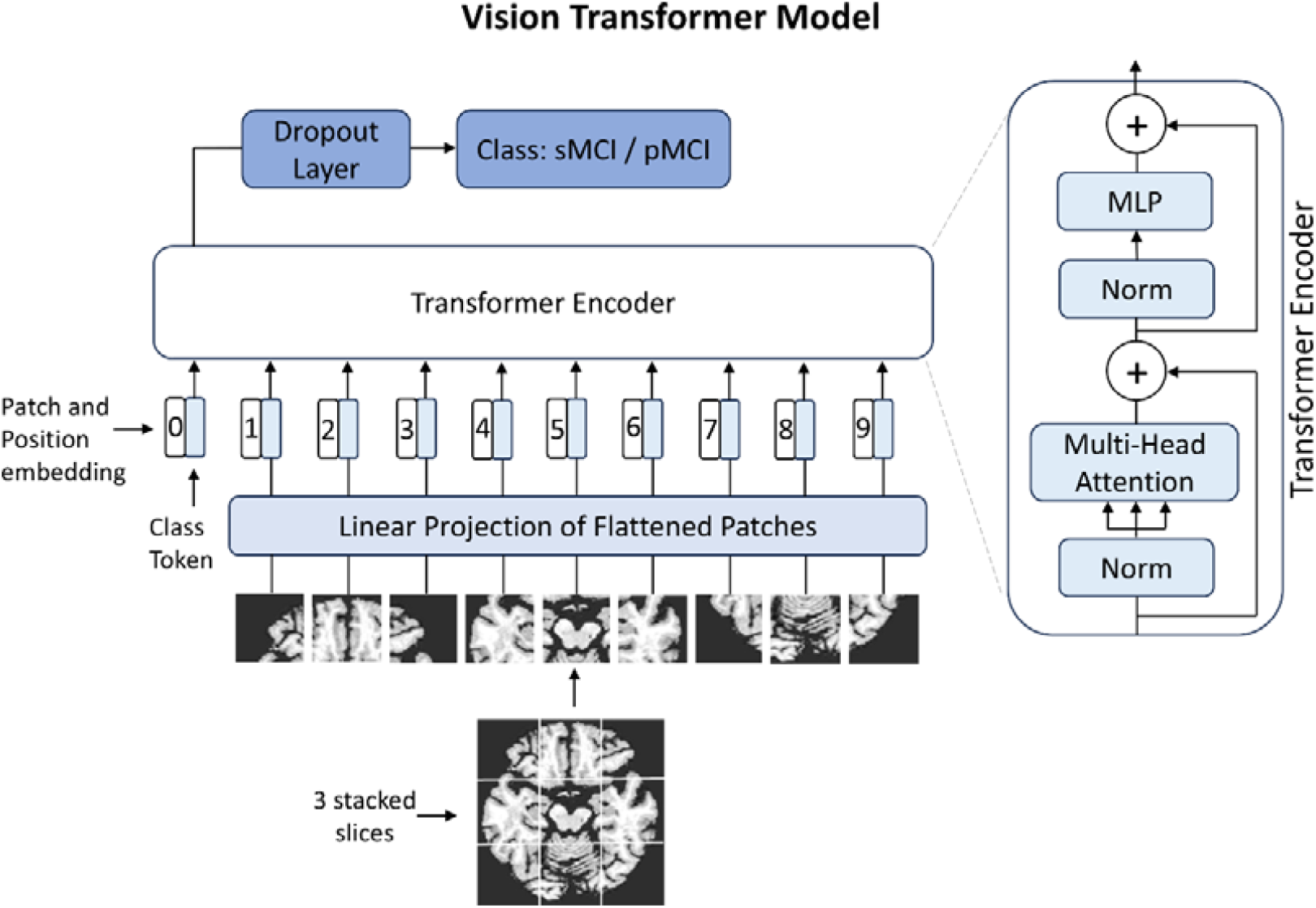
Model architecture featuring a customized MLP (multilayer perceptron) head with a dropout layer, adapted for binary classification. The backbone consists of 12 transformer encoder blocks and 12 multi-head attention modules, as in the pretrained ViT-base model. For illustration purposes, only 9 image patches are shown; the actual model processes 196 input patches. Architecture adapted from [5].

The three axial MRI slices of each subject were resized to 224 × 224 pixels to match the ViT’s input dimension. To account for intensity heterogeneity across the MRI scans, each subject’s three-slice volume was first normalized to a [0, 1] range using min-max scaling. Subsequently, the data was linearly transformed to the [-1, 1] range. This was achieved by subtracting 0.5 from each value and then dividing by 0.5, a common practice for fine-tuning.

### 2.5. Model Training and Fine-Tuning

The model was trained using PyTorch in a Google Colab environment with an NVIDIA A100 GPU (40 GB memory), 83.5 GB of system RAM. The data were split into training (70%), validation (15%), and test (15%) sets, while maintaining a constant sMCI/pMCI subject ratio across all splits. As data leakage is a serious issue frequently encountered in deep learning studies [18], data of each subject was assigned only once to one of the three folds to prevent overly optimistic and biased metrics. Data augmentation was applied during training, including random rotation (±10°) and horizontal flipping. Training was conducted using binary cross-entropy loss with logits, optimized using the AdamW optimizer (learning rate = 1e-5, weight decay = 0.05). The sigmoid function was applied to model outputs at inference time to obtain probability scores for binary classification. The model was trained for 30 epochs with a batch size of 64, and the best model was selected based on the epoch with the lowest validation loss. This process was repeated using various hyperparameter configurations as part of model tuning, with the best results achieved using the settings described above. The final performance of the model was evaluated on the independent test data set. A classification report was generated using the scikit-learn library, reporting the area under the receiver operating characteristic curve (ROC-AUC), accuracy, sensitivity, specificity, and F1-score.

## 3. Results

The comparison between the two groups regarding demographics showed no significant difference in gender distribution (Chi-square test, p = 0.58). Although the pMCI group was only 1.5 years older than the sMCI group, this age difference reached statistical significance (T-test, p = 0.013). Furthermore, both Mini-Mental State Examination (MMSE) and Clinical Dementia Rating Scale – Sum of Boxes (CDR-SOB) scores differed significantly (p < 0.001) between groups, with the pMCI cohort demonstrating poorer performance. Detailed results can be found in Table 1.

The distribution of conversion times of the pMCI group is depicted in Figure 2.

**Figure 2.**
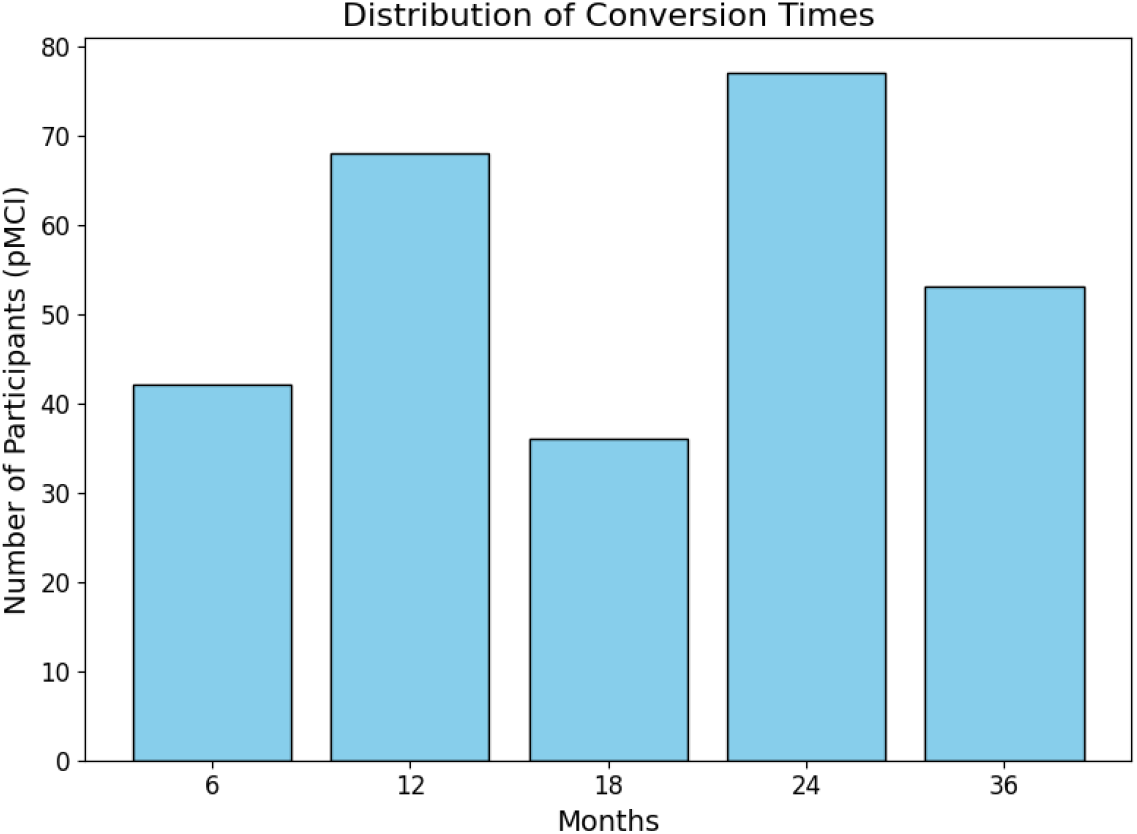
Overview of the distribution of conversion times in the pMCI group. Assessment frequency and intervals varied slightly between participants and across the different ADNI studies, and not all assessment periods were available in every study.

To obtain robust performance estimates and account for stochastic variability introduced by random weight initialization, data shuffling, and dropout, the model was trained and evaluated across 10 independent runs. On average, the model achieved an AUC-ROC of 0.74 with a standard deviation of 0.02 and a range of 0.72 to 0.78, indicating acceptable discriminative ability between the sMCI and pMCI groups. The mean accuracy was 0.69 (SD = 0.03; range = 0.63–0.74), sensitivity was 0.65 (SD = 0.07; range = 0.55–0.74), specificity was 0.72 (SD = 0.06; range = 0.58–0.78), and the F1-score for the pMCI class was 0.67 (SD = 0.04; range = 0.59–0.72).

Figure 3 shows the distribution of these performance metrics across all runs. The mean ROC and individual ROC curves and corresponding aggregated confusion matrix are displayed in Figure 4.

**Figure 3.**
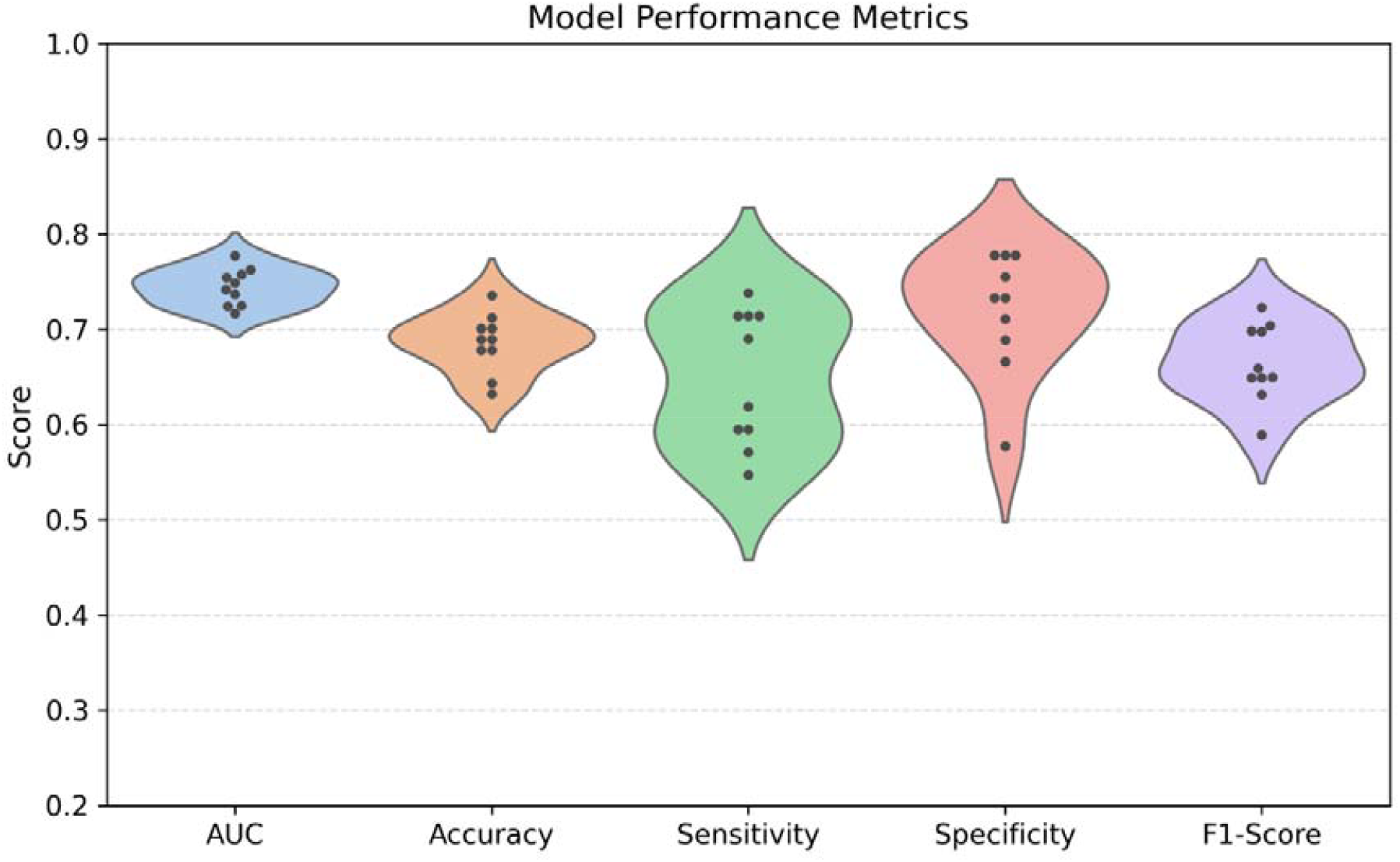
Distribution of model performance metrics across 10 runs of the test set (Area under the Curve (AUC), accuracy, sensitivity, specificity, and F1-score).

**Figure 4.**
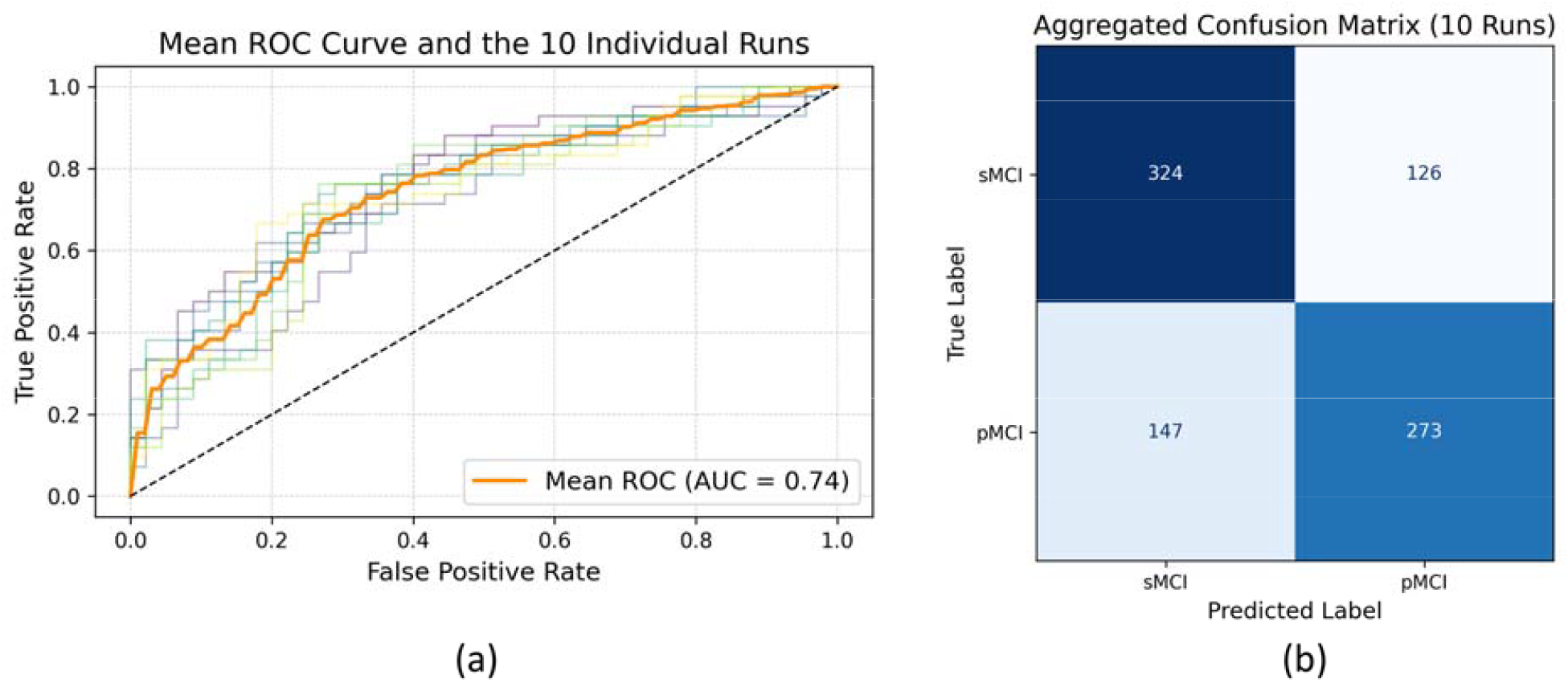
(a) Mean Receiver Operating Characteristic (ROC) curve (orange), including all 10 individual runs on the test dataset. Colored curves represent individual ROC curves from each run, illustrating performance variability. (b) Confusion matrix showing aggregated results from 10 independent model runs on the test dataset. Total counts of true and predicted labels for the sMCI and pMCI classes are summarized.

## 4. Discussion

In this investigation, we utilized axial T1-weighted MRI brain slices centered on the hippocampal region as input to a pretrained ViT model to predict conversion from MCI to AD. As the hippocampus is one of the first brain regions affected by the disease, we focused our analysis on this area to capture potential early structural changes. Specifically, we mapped three consecutive slices per subject to the three input channels of the 2D ViT model. Currently, no pretrained ViT models for 3D medical imaging trained on large-scale datasets comparable to the scale and diversity of ImageNet exist, which would provide more comprehensive representations of volumetric data and potentially deeper insights into disease progression. However, developing such models poses significant computational and data-related challenges. Hence, establishing large-scale 3D pretrained models that can be fine-tuned for specific clinical tasks remains a promising direction. ViTs represent a relatively recent advancement in computer vision and can complement traditional CNN approaches or even serve as a viable alternative, delivering results on par with CNNs or even surpassing their performance on tasks related to pathology detection [19]. Despite their advantages, such as modeling global context and long-range dependencies, ViTs are computationally expensive due to their quadratic complexity with respect to input size and large datasets are needed to reach best performance [5].

Here, we focused on the implementation of a standard ViT based solely on MRI as input data. A prior study has investigated MCI-to-AD conversion using a pre-trained ViT model with mid-sagittal slices [14]. An accuracy of 83% was reported in distinguishing MCI converters from non-converters. Their accuracy was higher than ours, as we achieved a mean accuracy of 69%. However, the presented steps in their investigation suggest that data from subjects were present in both the training and the validation and test sets, indicating data leakage. Thus, their reported accuracy metrics should be interpreted with caution, as they are likely overly optimistic and may not generalize well. Several studies employing architectures other than ViTs have investigated the prediction of MCI progression to AD based on structural MRI. CNNs have been utilized in numerous studies [18,20–26]. Models combining CNNs with attention mechanisms have also been explored [27–30]. Additionally, hybrid models integrating CNNs with transformer-based architectures have been proposed, including Hu et al. [31] with a Swin Transformer, Cao et al. [32] with MobileViT, and Khatri et al. [33]. Some of these studies reported high accuracies, with Ren et al. [28] achieving 87% and Khatri et al. [33] even exceeding the 90% mark. In particular, the latter also employed a ViT as a baseline for evaluating their proposed algorithm. They trained a ViT from scratch, achieving an accuracy of 79% for the MCI-to-AD prediction task. Notably, their comparison with other models showed that a ConViT [34] reached 85% accuracy, while a novel convolution-driven ViT model introduced by the authors achieved 91%. All models in their investigation were trained from scratch rather than leveraging pretrained weights, a choice that fundamentally limits the reliability of the reported results given their small dataset size. Furthermore, insufficient transparency regarding key methodological details complicates the interpretation of the reported accuracies in their study. Given the diversity in methodological approaches, specific techniques, and architectures across studies, a direct comparison of accuracy values does not reliably reflect actual model performance. More importantly, a specific algorithm must generalize well to new and unseen data and uses sound procedures for validation and to prevent any form of data leakage. This was systematically tested by [18], who applied various CNN architectures leading to balanced accuracy values ranging from 0.69 to 0.74 for the sMCI vs. pMCI classification task, which is comparable to our results. Future research should emphasize transparent validation protocols and explore hybrid architectures that integrate the strengths of both CNNs and transformers. Compared to other studies in the field, the ViT model proposed here falls within the range of reported metrics; however, it performs not as good as the best models discussed. As already mentioned, data leakage, particularly in studies involving slice-level data, is a well-documented issue in deep learning studies. This occurs when data from the same subject appears in both training and evaluation sets, allowing the model to memorize rather than generalize and thereby inflating performance estimates. While awareness of this problem has grown in recent years, it can still occur—particularly in studies with limited methodological transparency. Strict subject-level separation was maintained during data splitting to prevent any form of data leakage, as our dataset contained only one scan per subject.

In our investigation, we used only baseline MRI scans to train and evaluate our model. This decision reflects realistic clinical scenarios, where prognosis must be based on a single timepoint rather than longitudinal information. Including additional scans acquired closer to or even after conversion in progressive MCI individuals would have risked biasing the model toward easier cases and inflating performance, thereby limiting generalizability. Furthermore, we specified – as done in almost all related studies – a 36-month period for classifying individuals with MCI as progressive if they changed their status to AD within this period. However, individuals with stable MCI status were only included if they extended beyond that period and remained stable in all available follow-up assessments. This ensured that we did not label individuals as stable who may convert, e.g., shortly after the 36-month period, provided that assessment data were available. A strength of this study is the well-balanced dataset, which supports the reliability of reported metrics such as accuracy and F1-score as meaningful indicators of real-world model performance. While group differences in demographics and clinical scores were observed, these must be acknowledged as potential sources of bias. Specifically, the pMCI group was slightly older and performed worse on cognitive tests, which may have marginally influenced the results. However, the age difference was only 1.5 years on average, and while statistically significant, the effect is likely minimal. Nonetheless, this should be taken into account when interpreting the findings. Furthermore, as the ADNI study spans multiple sites and started already over a decade ago, the dataset includes images acquired using different scanners, field strengths, and acquisition protocols. Although this heterogeneity introduces variability, it also reflects real-world clinical diversity and may, in turn, enhance the generalizability of the model to broader populations. In addition, while MCI and AD statuses in ADNI were clinically defined without requiring biomarker confirmation, it should be noted that recent research is adopting a more biologically driven approach, defining Alzheimer’s disease as a continuum based on biomarker evidence [35].

Recent advances in model architectures, such as the combination of CNN and transformer architectures, show promising outcomes and represent a viable future direction as indicated above. New developments are already emerging, including revived state space models, in particular Mamba [36], which have been already applied to imaging tasks with Vision Mamba [37]. These models are not yet widely utilized in the medical domain but show promising potential also in the area of AD research.

In this work, we solely focused on MRI features, a choice that provides useful information while remaining relatively more accessible compared to other modalities, such as PET, where radioactive tracers are used. Furthermore, combining different input features with MRI, such as genetic and clinical data can also be seen as a viable approach [38]. Finally, we argue for the establishment of a global, standardized, unique and diverse test set, which could serve as a benchmark for future studies, allowing a fair comparison and realistic and unbiased estimation of each model’s performance. This would prevent biased results and limits the occurrence of overly optimistic accuracy metric reports.

## 5. Conclusions

Early prediction of whether a person with MCI will convert to AD or remain stable is still one of the greatest challenges in neuroscientific research. Given recent developments in the deep learning field with rapidly evolving model architectures, detecting early signs of progression, reflected in altered brain patterns based on structural MRI, is expected to become increasingly precise. Here, a standard ViT model was used, which is thought to capture complex and early patterns of brain atrophy, and was pretrained on large datasets of millions of images. We used three axial MRI slices covering the hippocampal area as input for prediction. With our pretrained and fine-tuned model, we were able to classify sMCI from pMCI individuals with moderate accuracy. However, our results were not as strong as those reported in some other studies where CNNs were primarily applied. To date, due to computational constraints, standard ViT approaches face challenges in processing entire 3D MRI volumes, which can be seen as a drawback compared to recent developments in CNN-based methods, where full 3D inputs are already feasible. However, this may become possible in the near future, potentially enabling even better classification performance, as early brain atrophy patterns are subtle and affect multiple brain regions in a complex manner.

## Author Contributions

Conceptualization, R.S. and P.F.; methodology, R.S.; software, R.S.; resources, P.F.; data curation, R.S.; writing—original draft preparation, R.S.; writing—review and editing, R.S.; visualization, R.S.; funding acquisition, R.S. All authors have read and agreed to the published version of the manuscript.

## Institutional Review Board Statement

All data used in this study were obtained from the Alzheimer’s Disease Neuroimaging Initiative (ADNI) database (adni.loni.usc.edu). Procedures and measurements were conducted in accordance with the Declaration of Helsinki and approved by the relevant Institutional Review Boards.

## Informed Consent Statement

Written informed consent was obtained from all participants.

## Data Availability Statement

Data available upon reasonable request.

## Acknowledgments

We acknowledge the support of large language models (LLMs) for proofreading, stylistic refinements and code assistance.

